# Circulation of respiratory viruses during the COVID-19 pandemic in The Gambia

**DOI:** 10.1101/2022.08.08.22278521

**Authors:** S. Jarju, E. Senghore, H. Brotherton, A. Saidykhan, S. Jallow, E. Krubally, E. Sinjanka, M.N. Ndene, F. Bajo, M. Sanyang, B. Saidy, A Bah, K. Forrest, E. Clarke, U. D’Alessandro, E. Usuf, C. Cerami, A. Roca, B. Kampmann, T. I. de Silva

## Abstract

In many countries, non-pharmaceutical interventions to limit SARS-CoV-2 transmission resulted in significant reductions in other respiratory viruses. However, similar data from Africa are limited. We explored the extent to which viruses such as influenza and rhinovirus co-circulated with SARS-CoV-2 in The Gambia during the COVID-19 pandemic. Between April 2020 and March 2022, respiratory viruses were detected using RT-PCR in nasopharyngeal swabs from 1397 participants with influenza-like illness. Overall virus positivity was 44.2%, with prevalence higher in children <5 years (80%) compared to children aged 5-17 years (53.1%), adults aged 18-50 (39.5%) and >50 years (39.9%), p<0.0001. After SARS-CoV-2 (18.3%), rhinoviruses (10.5%) and influenza viruses (5.5%) were the most prevalent. SARS-CoV-2 positivity was lower in children <5 (4.3%) and 5-17 years (12.7%) than in adults aged 18-50 (19.3%) and >50 years (24.3%), p<0.0001. In contrast, rhinoviruses were most prevalent in children <5 years (28.7%), followed by children aged 5-17 (15.8%), adults aged 18-50 (8.3%) and >50 years (6.3%), p<0.0001. Four SARS-CoV-2 waves occurred, with 36.1%-52.4% SARS-CoV-2 positivity during peak months. Influenza infections were observed in both 2020 and 2021 during the rainy season as expected (peak positivity 16.4%-23.5%). Peaks of rhinovirus were asynchronous to the months when SARS-CoV-2 and influenza peaked.

## Introduction

Many countries reported a dramatic reduction in the circulation of common respiratory viruses such as influenza and respiratory syncytial virus (RSV) during the initial stages of the COVID-19 pandemic^1-3^. This was attributed to the multiple non-pharmaceutical interventions (NPIs) implemented to reduce SARS-CoV-2 transmission such as bans on travel and social interactions, school closures, and mandatory wearing of face masks. Many of these early studies are from high-income countries with temperate climates, where the transmission patterns and seasonality of influenza and other respiratory viruses are well established^1-3^. In West Africa, surveillance for respiratory viruses is limited and data on the circulation of influenza and other viruses during the COVID-19 pandemic scarce.

The Gambia reported its first COVID-19 case on the 17^th^ March 2020 in a traveller^4^. A state of public emergency was declared on 27^th^ March 2020: NPIs such as social distancing, borders closure, closure of schools, ban on public gatherings, a night-time curfew, and compulsory wearing of face masks in public places were implemented. No increase in SARS-CoV-2 cases were seen in The Gambia until June 2020, when the first wave of infections occurred^5^. The state of emergency was relaxed on the 17^th^ September 2020 and since then, NPIs have not been enforced. Nevertheless, three further SARS-CoV-2 waves have occurred in The Gambia up to March 2022^5^.

We conducted a surveillance of respiratory viruses, including SARS-CoV-2, between April 2020 and March 2022. Our objectives were to describe the extent to which other respiratory viruses circulated alongside SARS-CoV-2 in The Gambia during the study period, whether there were differences in the prevalence of SARS-CoV-2 and other respiratory viruses across age groups, and whether peaks of other viruses such as human rhinovirus occurred at the same time or asynchronously with SARS-CoV-2.

## Methods

### Study setting, participant recruitment and sample collection

The Gambia has a population of approximately 2.5 million, with more than half living in urban coastal areas^6^. The climate is subtropical with a short rainy season between June and October, and a long dry season for the rest of the year^7^. Average temperatures range between 23°C to 33°C during the rainy season, and between 18°C to 30°C during the dry season^7^. Annual peaks of influenza virus infection usually occur during the rainy season^8^.

Nasopharyngeal swabs (NPS) from two surveillance studies were used for this analysis. In the first study, influenza-like illness (ILI) surveillance samples were collected from adults and children of all ages with acute respiratory illnesses attending the outpatient medical services at the Medical Research Council Unit The Gambia at the London School of Hygiene and Tropical Medicine (MRCG) clinics in Fajara (urban, coastal) and Keneba (rural). Patients presenting with shortness of breath, cough, or history of fever of any severity within the last 10 days provided written informed consent prior to study recruitment. Sample collection was carried out between April 2020 and June 2021 in Fajara and between September 2020 and June 2021 in Keneba.

In the second study, samples were collected between January 2021 and March 2022 from a clinical trial on COVID-19 treatment conducted in urban and peri-urban areas (PaTS study: NCT04703608). Written informed consent was obtained from participants aged ≥5 years presenting with acute fever and cough, or three or more of the following signs or symptoms: fever, cough, general weakness/fatigue, headache, myalgia, sore throat, coryza, dyspnoea, anorexia/nausea/vomiting, diarrhoea, recent loss of smell, recent loss of taste, altered mental status.

Both studies were approved by the Gambia Government and MRCG joint ethics committee and the Research Ethics Committee at the London School of Hygiene and Tropical Medicine (LSHTM).

### Respiratory virus detection

Ribonucleic acid (RNA) was extracted from 200µL of universal transport medium from NPS using the QIAamp Viral RNA kit (QIAGEN, Germany) as per the manufacturer’s instructions. All samples were spiked with Phocine herpes virus (PhHV) prior to extraction as a quality control for RNA extraction efficiency.

SARS-CoV-2 was detected using a Reverse Transcriptase Polymerase Chain Reaction (RT-PCR), with either the Takara One Step PrimeScript III RT-PCR Kit (TaKaRa Bio Inc., Japan) or the SuperScript III Platinum One-Step qRT-PCR Kit (Invitrogen, USA). Primers and probes targeting SARS-CoV-2 E and N genes, along with RNase P and PhHV were used previously described ^4^. Samples were considered positive if all four gene targets amplified at a cycle threshold (Ct) of 37 or below. In addition, a viral multiplex RT-PCR assay was used as previously described^8^ to detect influenza A and B, respiratory syncytial virus (RSV) A and B, parainfluenza viruses 1-4, human metapneumovirus (HMPV), adenovirus, seasonal coronaviruses (229E, OC43, NL63) and human rhinovirus. All samples with a Ct of ≤37 and passed quality control measures were considered positive.

### Data analysis

Clinical data, demographics and RT-PCR results were entered on a REDCap database. All results and metadata were downloaded from the database and analysed using Graphpad Prism V9.1.2. Statistical comparisons between group proportions were carried out using the two-sided chi-squared or Fisher’s exact test. All data required to replicate the analyses presented are available in Supplementary Table 1.

## Results

Among the 1397 participants recruited between April 2020 and March 2022, 617 (44.2%) were positive for one or more respiratory viruses. Virus positivity was significantly higher in children <5 years (80.0%, 95% CI 71.9-86.9%) than in the other age groups: children aged 5-17 years (53.1%, 45.1-61.1%), adults aged 18-50 years (39.5%, 36.2-42.9%), and adults >50 years (39.9%, 34.0-46.1%), p<0.0001 (Table 1).

**Table 1.**
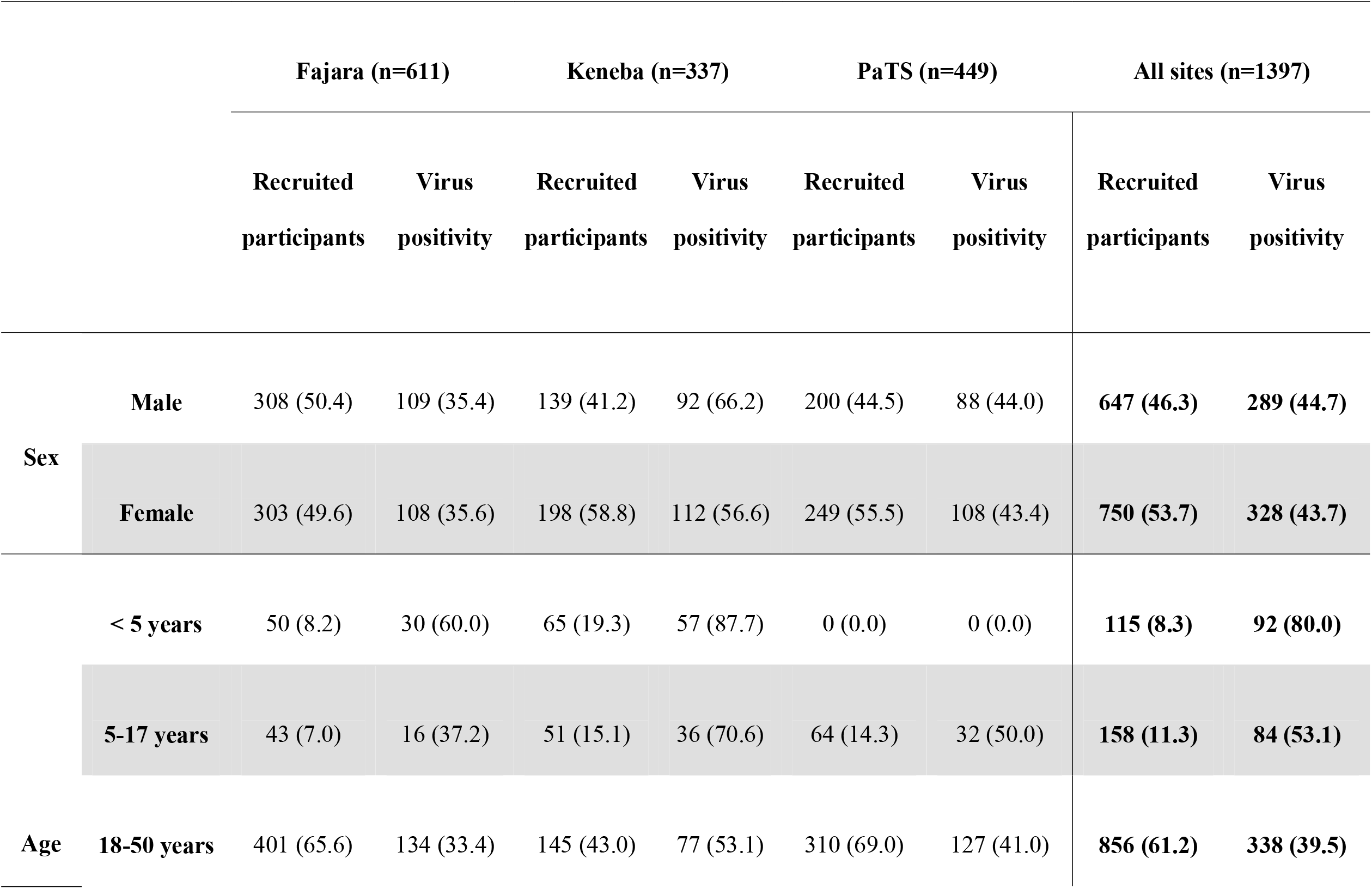
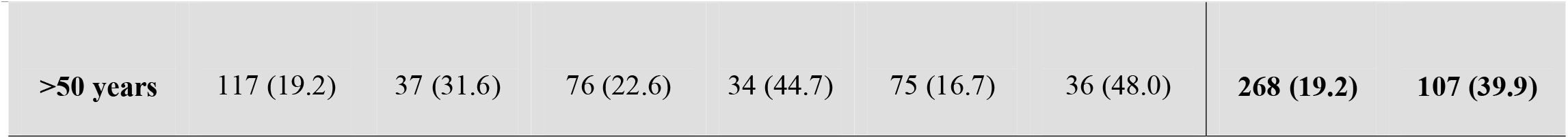
Recruited participants and percentage with respiratory viruses detected stratified by recruitment site, age and sex. Total numbers are shown with % in brackets. The denominators for age and sex percentages are the total recruited numbers at each site or all participants combined. Virus positivity denotes detection of one or more viruses in the sample. The denominators for respiratory virus positive % are the total recruited within each sex or age strata for each study or all participants combined. The PaTS trial did not recruit any children below the age of 5 years.

Overall, 255 (18.3%, 16.3-20.4) of samples tested during the study period were positive for SARS-CoV-2 (**Table 2**). The next most prevalent viruses were human rhinovirus (10.5%, 8.9-12.2%) and influenza A or B (5.5%, 4.4-6.8%). SARS-CoV-2 positivity was lower among children <5 years (4.3%, 1.4-9.9%) than in children aged 5-17 years (12.7%, 7.9-18.9%), adults aged 18-50 (19.3%, 16.7-22.1%), and adults >50 years (24.3%, 19.3-29.8%), p<0.0001. In contrast, human rhinovirus infections were most prevalent in children aged <5 years (28.7%, 20.7-37.9%), than in children aged 5-17 years (15.8%, 10.5-22.3%), adults aged 18-50 (8.3%, 6.5-10.4%), and adults >50 years (6.3%, 3.7-10.0%), p<0.0001. Most RSV A and HMPV infections were detected in children <5 years old, although the number of cases was relatively low (**Table 2**).

**Table 2.** Respiratory viruses detected during the study period stratified by age of participants. The denominator for virus positivity is total participants in the study (n=1397) or the number of participants recruited in each age group. RSV A = Respiratory Syncytial Virus A; HMPV = Human Metapneumovirus; HPIV = Human Parainfluenza viruses (pooled samples positive for HPIV1-4); Seasonal Coronavirus = combined 229E, NL63 and OC43 positive samples. Influenza A = samples positive for influenza A but negative in the pandemic H1N1 specific assay, which are presumed Influenza A H3N2, but not confirmed.

|  | **Virus** |  |  |  |  |
| --- | --- | --- | --- | --- | --- |
|  | **positivity** | **<5 years** | **5-17 years** | **18-50 years** | **>50 years** |
|  | **(%)** |  |  |  |  |
| **SARS-CoV-2** | 255 (18.3) | 5 (4.3) | 20 (12.7) | 165 (19.3) | 65 (24.3) |
| **Human Rhinovirus** | 146 (10.5) | 33 (28.7) | 25 (15.8) | 71 (8.3) | 17 (6.3) |
| **Influenza (all)** | 77 (5.5) | 10 (8.7) | 15 (9.5) | 42 (4.9) | 10 (3.7) |
| Influenza A | 29 (2.1) | 4 (3.5) | 7 (4.4) | 12 (1.4) | 6 (2.2) |
| Influenza A (H1N1) | 10 (0.7) | 0 (0.0) | 2 (1.3) | 8 (0.9) | 0 (0.0) |
| Influenza B | 38 (2.7) | 6 (5.2) | 6 (3.8) | 22 (2.6) | 4 (1.5) |
| **HPIV** | 47 (3.4) | 9 (7.8) | 8 (5.1) | 22 (2.6) | 8 (3.0) |
| **HMPV** | 38 (2.7) | 14 (12.2) | 9 (5.7) | 13 (1.5) | 2 (0.8) |
| **RSV A** | 25 (1.8) | 14(12.2) | 2 (1.3) | 9 (1.1) | 0 (0.0) |
| **Seasonal Coronavirus** | 26 (1.9) | 2 (1.7) | 4 (2.5) | 15 (1.8) | 5 (1.9) |
| **Adenovirus** | 2 (0.1) | 0 (0.0) | 1 (0.6) | 1 (0.1) | 0 (0.0) |
| **Samples tested** | 1397 | 115 | 158 | 856 | 268 |

Only 2.9% (41/1397) of recruited participants were hospitalized for their illness, although details of the clinical reasons for admission were not available. 15 (36.6%) of these admissions had respiratory viruses detected in NPS (3 RSV A, 3 Influenza B, 2 HMPV, 3 parainfluenza virus 1 and 4 human rhinovirus). 36.6% (15/41) of hospitalizations were in children <5 years old, representing a 13.0% (15/115) admission rate in this age group compared to 2.0% in the other age groups (26/1282), p<0.0001.

Four SARS-CoV-2 waves were observed in The Gambia during the 24-month study period (**Figure 1**); two during the rainy season (peak detection August 2020 and July 2021) and two during the dry season (peak detection March 2021 and January 2022). SARS-CoV-2 positivity during the four waves were 45.7%, 36.1%, 52.4% and 41.4%, respectively. Influenza A infections peaked in October 2020 (16.4% positivity) and October 2021 (23.5% positivity), with influenza B infections detected only in 2020 (peak November 2020, 23.3% positivity, **Figure 1**). Despite asynchronous peaks of SARS-CoV-2 and influenza, co-infections were observed, with 4/39 (10.3%) influenza A and 8/38 (21.1%) influenza B infected individuals also tested positive for SARS-CoV-2. 12.0% (3/25) of RSV A infections, 10.5% (4/38) of HMPV infections, 6.4% (3/47) of parainfluenza virus infections, and 4.1% (6/146) of human rhinovirus infections were also positive for SARS-CoV-2. Human rhinovirus positivity usually peaked at different times than SARS-CoV-2 and influenza (**Figure 1**).

**Figure 1.**
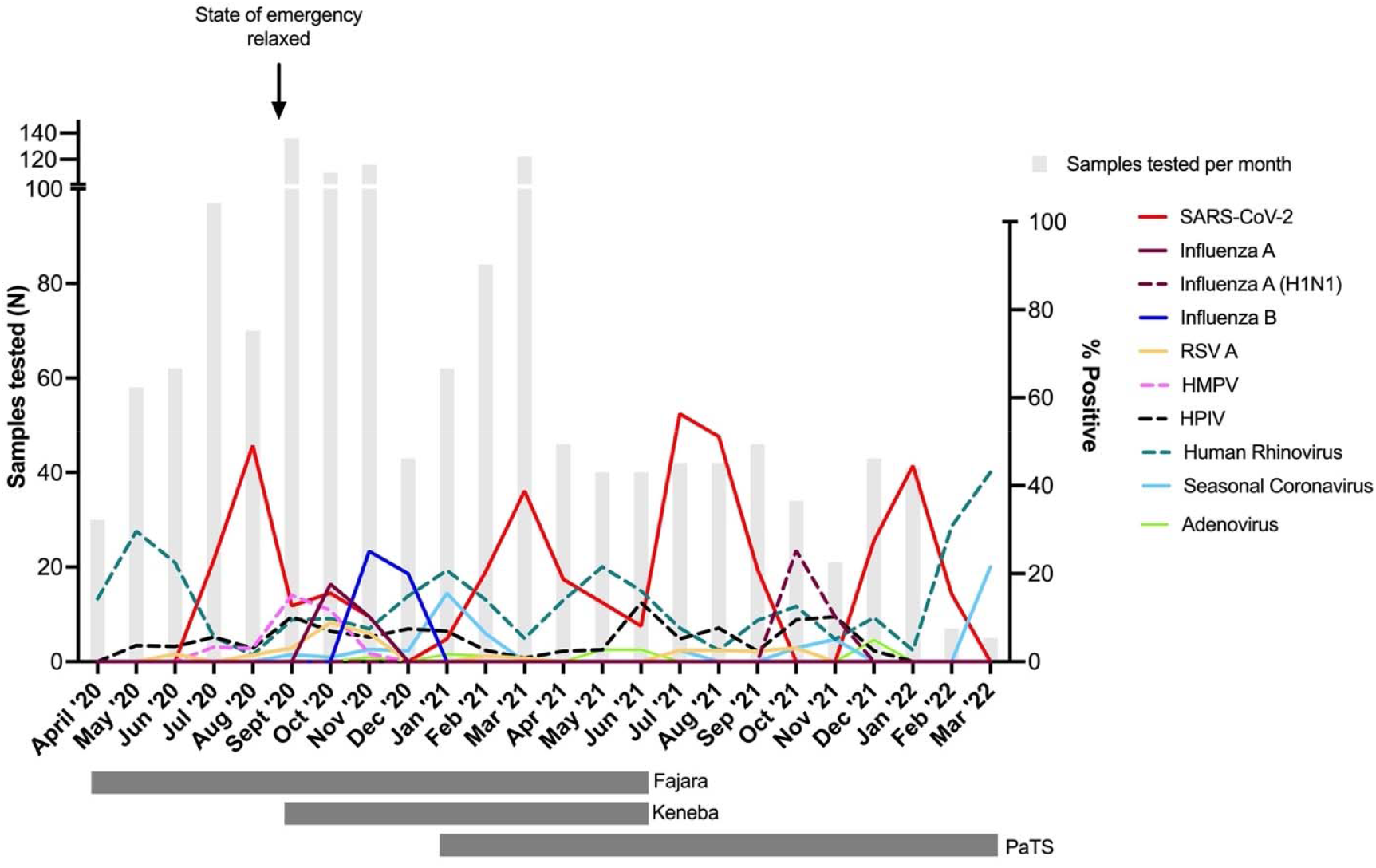
Percentage of influenza-like illness cases positive for respiratory viruses in The Gambia during April 2020 – March 2022. Vertical grey bars indicate the number of samples tested per month. Date when COVID-19 pandemic state of emergency was lifted is shown, after which time no restrictions or non-pharmaceutical interventions were in place within the country. Horizontal grey bars under the graph show periods when sample collection was done in the three participating sites (Fajara, Keneba, PaTS Study). Surveillance in Fajara was carried out between April 2020 and June 2021. Surveillance in Keneba was carried out between September 2020 and June 2021. Sample collection in the PaTS study was between January 2021 and March 2022. RSV A = Respiratory Syncytial Virus A; HMPV = Human Metapneumovirus; HPIV = Human Parainfluenza viruses (pooled samples positive for HPIV1-4); Seasonal Coronavirus = combined 229E, NL63 and OC43 positive samples. Influenza A = samples positive for influenza A but negative in the pandemic H1N1 specific assay.

## Discussion

SARS-CoV-2 was the most prevalent respiratory virus found among patients presenting with ILI during the COVID-19 pandemic in The Gambia. Although the seasonality of respiratory viruses is less predictable in tropical and sub-tropical than in temperate regions, many viral infections including influenza peak during the rainy season in The Gambia and neighbouring Senegal^8,9^. It is still unclear whether SARS-CoV-2 transmission will become seasonal, but The Gambia has experienced waves of infection in both the rainy and the dry season so far (twice per year), likely reflecting the emergence of new variants globally. Of note, the SARS-CoV-2 dynamics we have observed occurred in the context of no NPIs after the first wave and only 13.1% of the Gambian population fully vaccinated by February 2022^10^.

We observed peaks in influenza activity at expected times during 2020 and 2021, although without robust surveillance data from previous years, it is difficult to determine whether there was a reduction in transmission. Our previous study in children <5 years old with ILI attending outpatient services in Fajara found 8.4% influenza A or B positivity over 12 months in 2018-19^8^, compared to 7.3% in our current study which included both adults and children. In Senegal, between 1996 and 2009, 10.4% samples were positive for influenza A and 2.9% for influenza B^9^. Even in the absence of formal NPIs, either behavioural changes or reduction in global influenza transmission leading to reduced introductions could have an impact on influenza infections in The Gambia. However, the dramatic reductions in influenza seen in many high-income countries with strictly enforced NPIs do not appear to have occurred in The Gambia^1-3^. This may in part be due to poor adherence to NPIs, even when these measures were in place. In Canada, the average weekly positivity rate for influenza A in 2020-2021 was 0.012% compared to 10.40% in pre-pandemic years, and 0.006% for influenza B compared to 2.60% in pre-pandemic years^1^. Data from other African countries are scarce, with influenza and RSV reductions observed in South Africa^11^, but not in the Democratic Republic of the Congo (5.6% influenza A positivity and 0.9% influenza B positivity Dec 2020 – March 2021)^12^.

After SARS-CoV-2, human rhinovirus was the most prevalent virus detected in our samples. As expected, SARS-CoV-2 was responsible for a lower proportion of symptomatic ILI syndromes in children compared to adults, and the reverse pattern was seen for rhinovirus infections with decreasing prevalence with increasing age. Persistent circulation of human rhinoviruses has been described during the COVID-19 pandemic even in countries with NPI-driven reductions in other respiratory viruses, especially in the context of the reopening of schools^1,13^. This was also evident in our data with rhinoviruses circulating during the only period of NPI enforced in The Gambia, although schools were closed during that period. The high prevalence of human rhinovirus infections in children and the resulting increases in upper respiratory tract infections have been proposed as a potential mechanism for the reduced severity of COVID-19 observed in children^14,15^. The shared mucosal niche provides the opportunity for viral-viral interactions and interference between influenza and human rhinoviruses has been demonstrated in both epidemiological and *in vitro* data^16,17^. Whether similar interference between rhinoviruses and SARS-CoV-2 occurs to a degree population level transmission dynamics of either virus are affected is currently unclear. Our data on rhinovirus peaks and troughs during the study period should be treated with caution due to the small sample size and lack of pre-pandemic annual rhinovirus circulation patterns. Nevertheless, our findings do suggest that asynchronous circulation of SARS-CoV-2 and rhinoviruses may occur, supporting the experimental data from recent studies^14,15^. On the other hand, there is a further limitation to interpreting our prevalence data as it comes from two different studies with slightly different inclusion criteria.

In summary, our data show that many respiratory viruses continued to circulate during the COVID-19 pandemic in The Gambia, including human rhinoviruses, despite the presence of NPIs during the early stages of the pandemic, and influenza peaks during expected months. The age distributions of SARS-CoV-2 and human rhinovirus prevalence in symptomatic ILI are diametrically opposite, with rhinovirus infections driving the greater respiratory virus positivity seen in young children. The degree to which virus-virus interactions shape the impact of future SARS-CoV-2 waves and seasonality is important to establish as the transition to endemicity occurs in the coming years.

## Data Availability

All data produced in the present work are contained in the manuscript

## Acknowledgement

We would like to acknowledge all participants and their families who agreed to take part in the study. We would also like to acknowledge the contributions from Chiquita Jones, Mary Grey Johnson, Catherine Sarr and Fatoumatta Sillah in recruiting participants to the PaTS trial.

## Authors contribution

TdS, SJ, BK and AR conceived the study. SJ, AR, TdS, BK, HB, CC, KF, EC, UDA and EU were involved in study set up. ES, SJA, EK, NNM, FB and MS were involved in participants recruitment. SJ, ES, AS and AS were involved in laboratory processing. SJ and TdS analysed data. SJ, TdS, CC, BK and AR wrote the initial manuscript. All authors reviewed and edited the final manuscript.

